# The oligogenic structure of amyotrophic lateral sclerosis has genetic testing, counselling, and therapeutic implications

**DOI:** 10.1101/2024.03.21.24304693

**Authors:** Alfredo Iacoangeli, Allison A. Dilliott, Ahmad Al Khleifat, Peter M Andersen, Nazli A. Başak, Johnathan Cooper-Knock, Philippe Corcia, Philippe Couratier, Mamede de Carvalho, Vivian Drory, Jonathan D. Glass, Marc Gotkine, Yossef Lerner, Orla Hardiman, John E. Landers, Russell McLaughlin, Jesús S. Mora Pardina, Karen E. Morrison, Susana Pinto, Monica Povedano, Christopher E. Shaw, Pamela J. Shaw, Vincenzo Silani, Nicola Ticozzi, Philip Van Damme, Leonard H. van den Berg, Patrick Vourc’h, Markus Weber, Jan H. Veldink, Project MinE ALS Sequencing Consortium, Richard JB Dobson, Guy A. Rouleau, Ammar Al Chalabi, Sali M.K. Farhan

## Abstract

Recently, large-scale case-control analyses have been prioritized in the study of ALS. Yet the same effort has not been put forward to investigate additive moderate phenotypic effects of genetic variants in genes driving ALS risk, despite case-level evidence suggesting a potential oligogenic risk model. Considering its direct clinical and therapeutic implications, a large-scale robust investigation of oligogenicity in ALS is greatly needed.

Here, we leveraged the Project MinE ALS Sequencing Consortium genome sequencing datasets of individuals with ALS (n = 6711) and controls (n = 2391) to identify signals of association between oligogenicity in known ALS genes (n=26) and disease risk, as well as clinical outcomes.

Applying regression models to a discovery and replication cohort, we observed that the risk imparted from carrying rare variants in multiple known ALS genes was significant and was greater than the risk associated with carrying only a single rare variant, both in the presence and absence of variants in the most well-established ALS genes, such as *C9orf72*. However, in contrast to risk, the relationships between oligogenicity and ALS clinical outcomes, such as age of onset and survival, might not follow the same pattern as we did not observe any associations.

Our findings represent the first large-scale, case-control assessment of oligogenic associations in ALS to date and confirm that oligogenic events involving known ALS risk genes are indeed relevant for the risk of disease in approximately 6% of ALS but not necessarily for disease onset and survival. This must be considered in genetic counselling and testing by ensuring the use of comprehensive gene panels even when a potential pathogenic variant has already been identified. Moreover, in the age of stratified medication and gene therapy, it supports the need of a complete genetic profile for the correct choice of therapy in all ALS patients.

## Introduction

As an intermediary between monogenic and polygenic disease transmission, oligogenic inheritance refers to the additive moderate phenotypic effect of genetic variants in a few genes that together drive disease presentation. Establishing whether oligogenicity plays a role in the development of a disease is important for more accurate diagnosis, as it may clarify the role of low penetrance variants or explain atypical clinical presentations. It could also drive the development of treatment by highlighting multiple potential targets. Therefore, identifying an oligogenic component of risk in a disease considered to be predominantly monogenic has direct implication for genetic counseling and risk assessment^1^. Although not well studied, oligogenicity has been described in a selected number of diseases such as Bardet-Biedl syndrome, Charcot-Marie-Tooth (CMT), and long QT syndrome^2–8^. Interestingly, all three forms of transmission — monogenic, polygenic and oligogenic — have been reported in amyotrophic lateral sclerosis (ALS).

In recent years, large-scale case-control analyses have been prioritized in the study of monogenic and polygenic forms of ALS, often including thousands of samples to maximize the statistical power of discovery^9–16^. However, the same effort has not been put forward to investigate oligogenic events driving ALS risk. Case-level evidence has suggested an oligogenic risk model in ALS^17,18^, which conforms with the proposed multistep hypothesis of ALS that describes multiple molecular events — including the possibility of multiple genetic variants — occurring to trigger ALS onset^19,20^. Yet large case-control analyses have not been performed using ALS cohorts to confirm the role of oligogenicity in the disease. Previous reports of the phenomenon had modest sample sizes, or were limited to case studies that did not include control cohorts, and did not directly assess the disease risk or influence on clinical outcomes imparted by carrying multiple variants in ALS genes with respect to carrying only one^17,18,21–27^.

Here, we leveraged the Project MinE ALS Sequencing Consortium large-scale, genome sequencing datasets of individuals with ALS (n = 6711) and controls (n = 2391) to identify signals of association between oligogenicity in known ALS genes and disease risk. Further, we investigated whether carrying multiple rare variants influences clinical features of disease, including age of onset and survival. The presented analyses represent the first large-scale, case-control assessment of oligogenic associations in ALS to date.

## Methods

### Study cohort and sample sequencing

Genome sequencing data obtained from the Project MinE ALS Sequencing Consortium were comprised of a discovery subset (individuals with ALS = 4518 and controls = 1821) and a replication cohort (individuals with ALS = 2193 and controls = 570) corresponding to the two main releases of Project MinE (data freeze 1 and 2). Details regarding sequencing methodology and quality control were previously described^28,29^. Briefly, all samples were sequenced using either the Illumina HiSeq 2000 platform or Illumina HighSeq X platform (San Diego, CA, USA), and sequencing reads were aligned to the hg19 reference genome to call single nucleotide variants, insertions, and deletions. Quality control was performed at both an individual and variant level, and included assessment of read depth and coverage, ancestry-defining principal component analysis (PCA), and identity-by-descent analysis. Controls were matched to the individuals with ALS based on age, sex, and geographical region.

Post quality control, 4299 individuals with ALS and 1815 controls from the Project MinE discovery subset, and 2057 individuals with ALS and 513 controls from the replication cohort were retained for analysis. Genetic data and clinical outcomes, including age of ALS onset and survival period, were available for the ALS patients. Survival period was defined as years from diagnosis to death, or years from diagnosis to last follow-up, as appropriate.

### Variant annotation and filtering

Variants were annotated using the Ensembl variant effect predictor (VEP) and Annovar^30,31^. Variants were further annotated based on minor allele frequencies (MAFs) using the Genome Aggregation Database (gnomAD) v2.1.1 non-neurological dataset^32^. Only variants considered rare, with an MAF < 0.01 in both gnomAD and the Project MinE controls, were retained for further analysis.

Twenty-six genes previously associated with ALS in the literature were selected for the analysis, as previously described^33^, and are hereafter referred to as known ALS genes (**Supplemental Table 1**). Only genes with at least two publications reporting rare coding variants in individuals with ALS were included. Genes with unclear inheritance patterns or limited or refuted gene-disease validity classification from the ClinGen ALS Gene Curation Expert Panel (GCEP) as of January 2022 were excluded (<u>Clinical Genome ALS</u>). Genes were also subclassified based on the strength of evidence regarding their association with ALS. “Established ALS genes” included those with a definitive relationship with ALS based on curation by the ClinGen ALS GCEP. All other genes were considered “ALS-associated genes”. For *C9orf72* and *ATXN2* only the ALS-associated repeat expansions were included, while for the remaining 24 known ALS genes only variants within the protein coding regions were retained.

The rare, protein-coding variants were binned into three classes: 1) synonymous variants; 2) missense variants (missense single nucleotide variants and in-frame insertions or deletions); and 3) protein truncating variants (PTVs; stop lost and gained, start lost, transcript amplification, frameshift, transcript ablation, and splice acceptor and donor variants).

### Pathogenic repeat expansion detection

All genome samples were also assessed for the *C9orf72* GGGGCC and *AXTN2* CAG repeat expansions using ExpansionHunter^34^. ExpansionHunter has been previously validated for the repeat expansions in both *C9orf72* and *ATXN2*^35–37^. A hexanucleotide repeat expansion of >30 copies in *C9orf72* is considered pathogenic for ALS, whereas a trinucleotide intermediate repeat expansion of 29-32 copies in *ATXN2* is considered a risk factor for ALS^38–40^.

### Statistical analyses

To model the influence of carrying a single variant in a known ALS gene (singleton) or carrying at least one variant in more than one known ALS genes (oligogenic) on ALS risk and clinical outcomes, regression analyses were applied. More specifically, risk of ALS as a function of singleton or oligogenic carrier status were assessed in individuals with ALS and controls using logistic regressions adjusting for sex, ten ancestry defining PCs, and total genetic load (summation of synonymous, missense, and PTVs, as previously described)^9^. Associations between ALS age of onset and singleton or oligogenic carrier status in individuals with ALS were assessed using linear regressions adjusting for sex, site of onset, ten ancestry defining PCs, and total genetic load. Finally, associations between ALS survival period and singleton or oligogenic carrier status in individuals with ALS were assessed using Cox proportional hazard regressions adjusting for the same co-variates described above.

To control for any residual synonymous inflation not fully corrected by population structure based PCs, and to determine whether the *C9orf72* or *ATXN2* repeat expansions were driving associations, all regression models assessed associations with five variant type bins: 1) synonymous variants; 2) missense variants and PTVs; 3) missense variants, PTVs, and *ATXN2* repeats; 4) missense variants, PTVs, and *C9orf72* repeats; and 5) missense variants, PTVs, *ATXN2* repeats and *C9orf72* repeats. Statistical analyses were performed using the *logistf* library from R statistical software [4.2.2]^41^ in R Studio [1.1.414]. Data visualization was performed using the *ggplot2* R package (v3.3.6)^42^.

## Results

### Oligogenic risk of ALS

Following the assessment for rare single nucleotide variants and repeat expansions in known ALS genes, we determined the number of singleton and oligogenic carriers (Table 1). In total, 1161 individuals with ALS (n = 4299) were considered singleton carriers, defined as carrying one non-synonymous, rare variant in one known ALS gene, in the Project MinE discovery subset. Whereas 255 individuals with ALS were considered oligogenic carriers, defined as carrying at least one non-synonymous, rare variant in two or more known ALS genes, in the Project MinE discovery subset. Similarly, 540 individuals with ALS (n = 2057) and 131 individuals with ALS were considered singleton and oligogenic carriers, respectively in the Project MinE replication subset.

**Table 1.** Age of onset and survival based on the number and type of rare variants (MAF < 0.01) carried by individuals with ALS from the Project MinE ALS Sequencing Consortium.

| Variant Type | ALS Carriers<br>(n = 4299) | Age of Onset (years) |  |  | Survival (years) |  |  |
| --- | --- | --- | --- | --- | --- | --- | --- |
|  |  | Mean | Median | SD | Mean | Median | SD |
| No Variants | 2883 | 62.67 | 63.57 | 11.71 | 3.42 | 2.71 | 2.65 |
| Singleton | 1161 | 61.60 | 62.65 | 11.50 | 3.43 | 2.70 | 2.71 |
| Oligogenic (All ALS Genes) | 255 | 61.83 | 62.90 | 10.93 | 3.47 | 2.71 | 3.02 |
| Oligogenic (Established ALS Genes) | 129 | 60.85 | 61.57 | 10.63 | 3.68 | 2.87 | 3.09 |
| Oligogenic (ALS-Associated Genes) | 31 | 62.24 | 65.70 | 11.90 | 2.93 | 2.25 | 2.16 |
| Oligogenic ($\geq 1$ Variant in Established ALS Gene & $\geq 1$ Variant in ALS-Associated Gene) | 95 | 63.41 | 64.94 | 10.93 | 3.49 | 2.65 | 3.28 |

Minor allele frequencies were obtained from the GnomAD v2.1.1 non-neurological dataset. Singleton refers to carrying one variant in one ALS gene. Oligogenic refers to carrying at least one variant in $\geq 2$ ALS genes. Established ALS genes include those with a definitive relationship with ALS based on curation by the ClinGen ALS Gene Curation Expert Panel. All other genes were considered ALS-associated genes.

Using our model, an enrichment analysis indeed demonstrated that carrying ≥ 2 rare (MAF < 0.01), non-synonymous variants in multiple ALS genes was significantly associated with disease risk, with greater odds than observed in the singleton analysis (**Figure 1**). The increased odds relative to singleton carrier status was observed for oligogenic carriers when only missense variants or PTVs were considered (singleton OR = 1.22 [1.09-1.37], p = 1.77e-03; oligogenic OR = 1.46 [1.20-1.78], p = 1.70e-04), as well as when *C9orf72* repeat expansions (singleton OR = 1.42 [1.34-1.51], p = 3.21e-08; oligogenic OR = 1.82 [1.50-2.22], p = 4.99e-10), *AXTN2* repeat expansions (singleton OR = 1.22 [1.09-1.37], p = 2.50e-03; oligogenic OR = 1.65 [1.23-2.21], p = 5.00e-04), and both repeat expansions (singleton OR = 1.49 [1.30-1.71], p = 1.22e-09; oligogenic OR = 2.27 [1.69-3.05], p = 1.70e-09) were included in the analysis.

**Figure 1.**
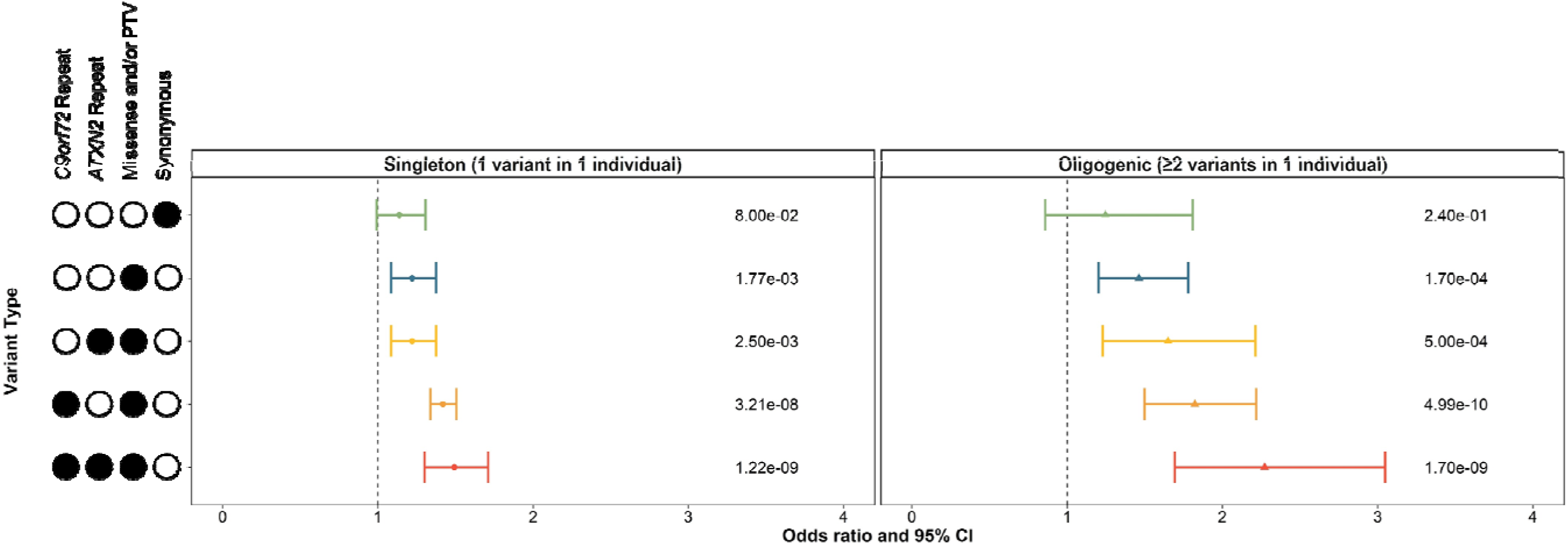
Singleton and oligogenic enrichment of rare variants (MAF < 0.01) in known ALS genes in individuals with ALS compared to controls. Enrichment of carriers of rare variants in one known ALS gene and carriers of rare variants in two or more known ALS genes was compared to non-carriers in individuals with ALS compared to controls. Enrichment analyses were performed using logistic regression in the discovery subset (individuals with ALS = 4299, controls = 1815) of the Project MinE ALS sequencing consortium dataset, including sex, 10 ancestry defining principal components, and total variant count (total genetic load) as covariates. The upset plot on the left legend indicates the variant types that the singleton or oligogenic variant may encompass; for example, the bottom row indicates the singleton variant or two or more oligogenic variants may be a *C9orf72* repeat expansion, *ATXN2* repeat expansion, missense variants, or protein truncating variants (PTVs). Synonymous variants, missense variants, and PTVs were identified in 24 known ALS genes using whole genome sequencing. *ATXN2* and *C9orf72* refer to a pathogenic repeat expansion being identified in the respective gene using ExpansionHunter. Minor allele frequencies were obtained from the GnomAD v2.1.1 non-neurological dataset. Abbreviations: CI, confidence interval; MAF, minor allele frequency; PTV, protein truncating variant.

Upon confirmation of our risk analysis using the Project MinE replication subset, we observed that singleton carrier status was only significantly associated with ALS when missense variants or PTVs in known ALS genes and the *C9orf72* and *ATXN2* repeat expansions were included in the analysis (OR = 1.30 [1.05-1.61], p = 1.70e-02; **Supplemental Figure 1**). In contrast, oligogenic carrier status was significantly associated with ALS when only missense variants or PTVs were considered (OR = 1.39 [1.02-1.90], p = 4.00e-02), as well as when *C9orf72* repeat expansions (OR = 1.57 [1.15-2.15], p = 4.50e-03), *AXTN2* repeat expansions (OR = 1.49 [1.07-2.08], p = 1.30e-02), and both repeat expansions (OR = 1.73 [1.24-2.42], p = 5.70e-04) were included in the analysis.

We also assessed whether oligogenic carrier status of variants with lower MAFs were associated with an increased risk of ALS in comparison to singleton carrier status. In a similar manner to the MAF < 0.01 rare variant assessment, oligogenic carrier status for variants of MAF < 0.001, MAF < 0.0001, and AC = 1 were significantly associated with disease risk, with greater odds than observed in the singleton analyses of the Project MinE discovery subset (**Supplemental Figure 2**). These findings were replicated in the analyses of singleton and oligogenic carrier statuses of variants with lower MAFs in the Project MinE replication cohort (**Supplemental Figure 1**).

### Oligogenic influence on ALS clinical outcomes

From the Project MinE discovery cohort, 4299 individuals with ALS had their age of onset and survival periods captured. Summary of the clinical outcomes of individuals with ALS carrying zero, singleton, or oligogenic non-synonymous, rare variants in known ALS genes are presented in **Table 1**.

Linear regressions adjusting for sex, site of onset, ten ancestry defining PCs, and total variant count were applied to determine whether singleton or oligogenic carrier statuses were associated with ALS age of onset. We found that carrying a singleton, rare (MAF < 0.01), non-synonymous variant was significantly associated with lower age of onset when the *C9orf72* repeat expansion was included in the analysis (β = -1.40 [-2.24 - -0.56], p = 1.4e-03; **Figure 2**). The singleton association was also observed for variants with an MAF < 0.001 and MAF < 0.0001 when the *C9orf72* repeat expansion was included in the analyses and for MAF < 0.0001 when the *C9orf72* repeat expansion was excluded (p = 0.031) (**Supplemental Figure 3**).

**Figure 2.**
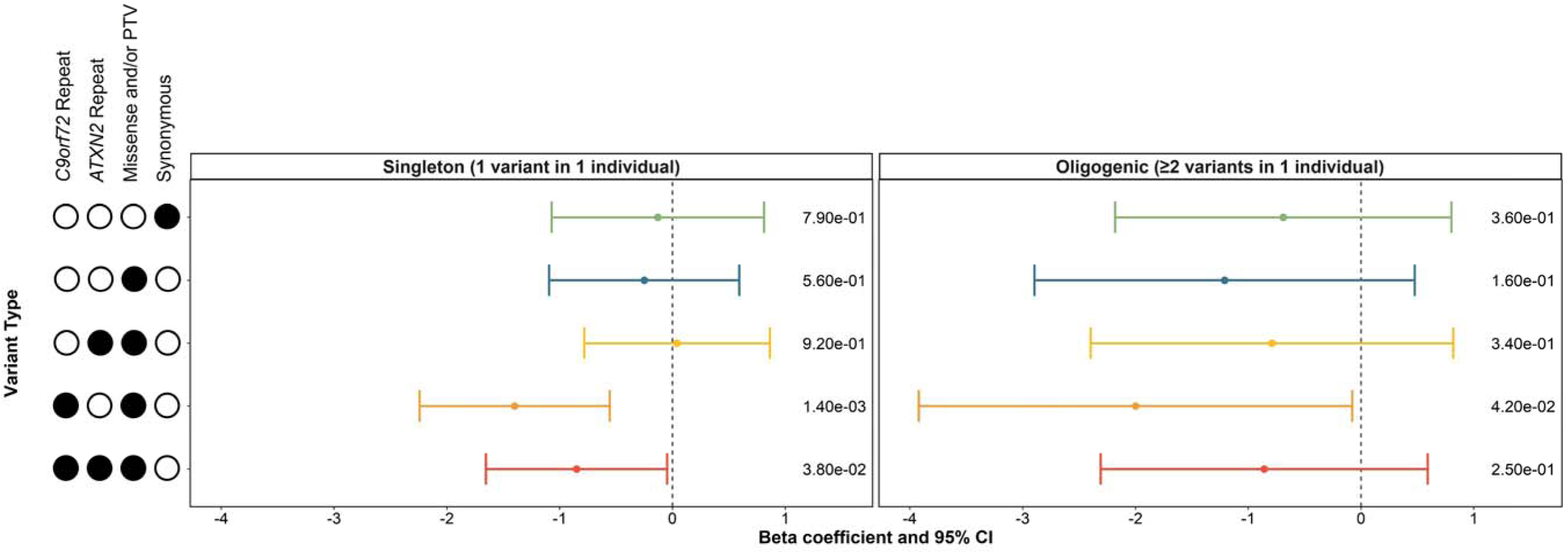
Influence of singleton and oligogenic enrichment of rare variants (MAF < 0.01) in known ALS genes to ALS age of onset. The influence of carrying a rare variant in a single known ALS genes was compared to the influence of carrying rare variants in two or more known ALS genes on ALS age of onset in the discovery cohort of the Project MinE ALS sequencing consortium (individuals with ALS = 4299). Enrichment analyses were performed using linear regression including sex, site of onset, 10 ancestry defining principal components, and total variant count (total genetic load) as covariates. The upset plot on the left legend indicates the variant types that the singleton or oligogenic variant may encompass; for example, the bottom row indicates the singleton variant or two or more oligogenic variants may be a *C9orf72* repeat expansion, *ATXN2* repeat expansion, missense variants, or protein truncating variants (PTVs). Synonymous variants, missense variants, and PTVs were identified in 24 known ALS genes using whole genome sequencing. *ATXN2* and *C9orf72* refer to a pathogenic repeat expansion being identified in the respective gene using ExpansionHunter. Minor allele frequencies were obtained from the GnomAD v2.1.1 non-neurological dataset. Abbreviations: CI, confidence interval; MAF, minor allele frequency; PTV, protein truncating variant.

Carrying oligogenic, rare (MAF < 0.01), non-synonymous variants in multiple known ALS genes was only marginally significantly associated with age of onset when the *C9orf72* repeat expansion was included in the analysis (p = 0.042, **Figure 2**), while a lack of oligogenic association was observed for variants of lower MAF (**Supplemental Figure 3**).

Similarly, Cox proportional hazard models adjusting for sex, site of onset, ten ancestry defining PCs, and total variant count, were applied to determine whether singleton or oligogenic carrier statuses were associated with ALS survival period. We found that carrying a singleton, rare (MAF < 0.01), non-synonymous variant was not significantly associated with ALS survival period (**Figure 3**); however, carrying a singleton missense variant or PTV of MAF < 0.001 and MAF < 0.0001, or a *C9orf72* repeat expansion were significantly associated with an increased hazard ratio (p_0.001_ = 0.041 and p_0.0001_ = 0.0056, **Supplemental Figure 4**).

**Figure 3.**
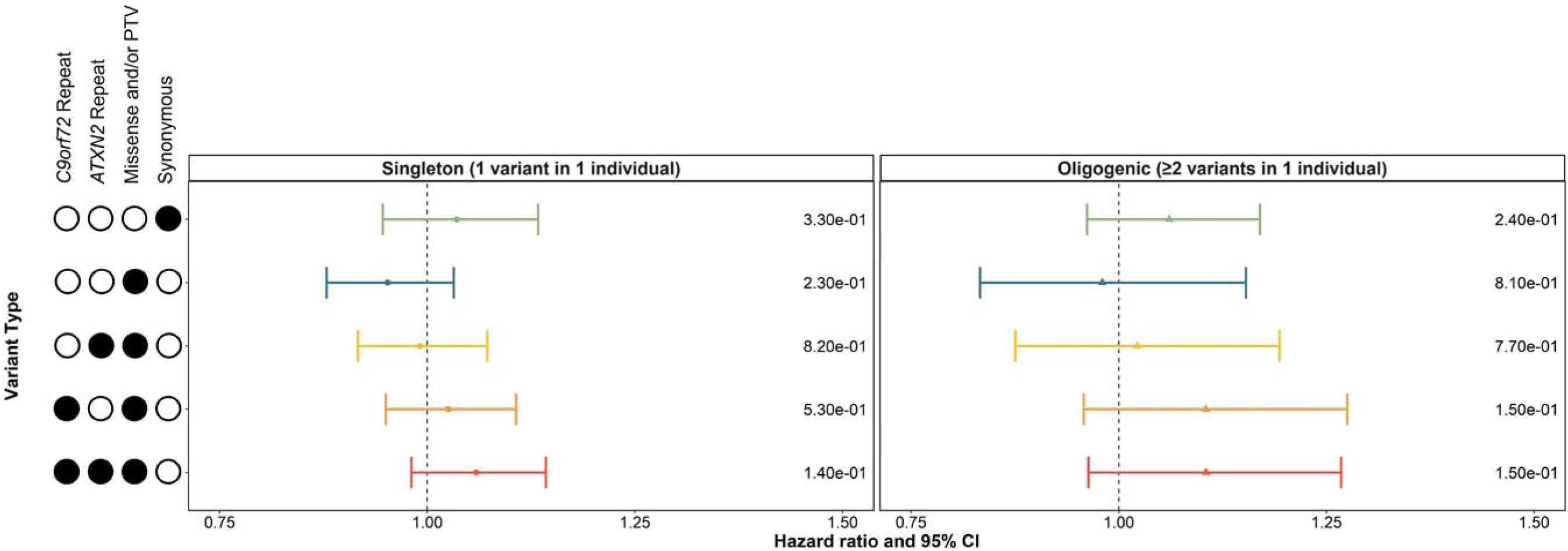
Influence of singleton and oligogenic enrichment of rare variants (MAF < 0.01) in known ALS genes to ALS survival period. The influence of carrying a rare variant in a single known ALS gene was compared to the influence of carrying rare variants in two or more known ALS genes on ALS survival period in the discovery cohort of the Project MinE ALS sequencing consortium (individuals with ALS = 4299). Enrichment analyses were performed using Cox proportional-hazards models including sex, site of onset, 10 ancestry defining principal components, and total variant count (total genetic load) as covariates. The upset plot on the left legend indicates the variant types that the singleto or oligogenic variant may encompass; for example, the bottom row indicates the singleton variant or two or more oligogenic variants may be a *C9orf72* repeat expansion, *ATXN2* repeat expansion, missense variants, or protein truncating variants (PTVs). Synonymous variants, missense variants, and PTVs were identified in 24 known ALS genes using whole genome sequencing. *ATXN2* and *C9orf72* refer to a pathogenic repeat expansion being identified in the respective gene using ExpansionHunter. Minor allele frequencies were obtained from the GnomAD v2.1.1 non-neurological dataset. Survival period was defined as years from diagnosis to death, or years from diagnosis to last follow-up, as appropriate. Abbreviations: CI, confidence interval; MAF, minor allele frequency; PTV, protein truncating variant.

Carrying oligogenic, rare, non-synonymous variants in multiple known ALS genes was not associated with decreased or increased survival period, for any MAF classes (**Supplemental Figure 4**).

### Gene involvement in oligogenic events

In the Project MinE discovery cohort, individuals with ALS had the highest frequency of oligogenic carriers when one of the rare variants carried was in *NEK1* (1.84%) closely followed by *ANXA11* (1.81%) and the *C9orf72* repeat expansion (1.48%; **Figure 4A,B**). However, *ANXA11* and *NEK1* were also the genes with the highest oligogenic carrier frequencies in controls (1.37% and 0.99%, respectively), whereas the *C9orf72* repeat expansion was only observed in one control with another rare variant in a known ALS gene, specifically *VAPB* (**Supplemental Figure 5-6**).

**Figure 4.**
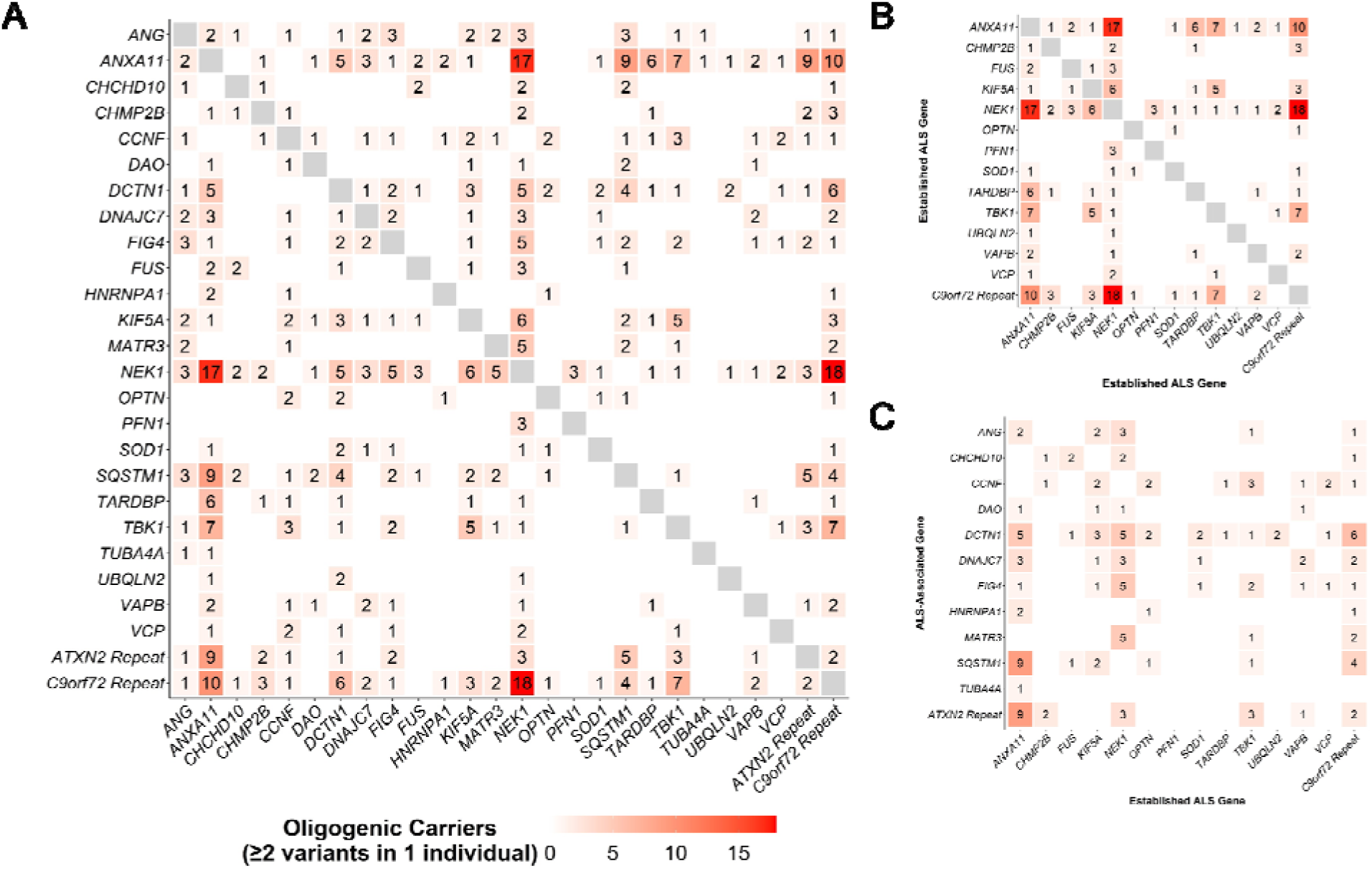
Individuals with ALS carrying two or more rare variants in ALS genes. **(A)** The number of individuals with ALS from the Project MinE discovery cohort (n = 4299) carrying at least one rare variant in each gene encompassed in the respective column and row. **(B)** The number of individuals with ALS from the Project MinE discovery cohort carrying at least one rare variant in an established ALS gene encompassed in the respective column and row. **(C)** The number of individuals with ALS from the Project MinE discovery cohort carrying at least one primary rare variant in an established ALS gene encompassed in the respective column and at least one secondary rare variant in a known ALS gene in the respective row. Established ALS genes were defined as those with a definitive ALS gene-disease relationship based on review by the ClinGen ALS Gene Curation Expert Panel. All remaining genes assessed were considered ALS-associated genes.

The only genes for which oligogenic carriers with ALS but no controls were observed carrying at least one rare variant in the gene along with a rare variant in another known ALS gene were *CHMP2B*, *PFN1*, *SOD1*, and *UBQLN2* (**Supplemental Figure 5**). Oligogenic carriers with ALS with two or more rare variants in the most well-established ALS genes are displayed in **Figure 4B**. Although these are considered very rare events, 129 carriers with ALS were observed across the Project MinE discovery cohort (2.8%). Oligogenic carriers with ALS with one or more rare variant(s) in a well-established ALS gene in addition to one or more rare variant(s) in an ALS-associated gene are displayed in **Figure 4C**.

## Discussion

While evidence from previous case reports has suggested that there could be an oligogenic burden to developing ALS, our study is the first at scale involving large discovery and replication datasets of thousands of people with ALS and controls. Across the two subsets of the Project MinE Sequencing Consortium, 6% of individuals with ALS were considered oligogenic carriers. This proportion was relatively comparable to the 6.82% of Australian individuals with sporadic ALS and 3.8% of North American individuals with ALS that were found to be oligogenic carriers in cohorts of more modest sample size^18,21^. In this oligogenic subgroup of people with ALS, we observed that the risk imparted from carrying rare variants in multiple known ALS genes was significant and was greater than the risk associated with carrying only a single rare variant, in principle consistent with the multi-step hypothesis of ALS^19,20^. Further, our results have direct implication for genetic counselling and testing. Having shown that variants in more than one known ALS gene affect risk in approximately 6% of patients, it follows that all ALS-associated genes must be included in clinical genetic testing even when a potential pathogenic variant has already been identified. Without the use of comprehensive gene panels, complications may arise in cases of familial testing, whereby even in families for which there is a known pathogenic gene variant, the lack of knowledge regarding any additional variants contributing to disease risk may result in false reassurance in the case of a negative genetic test. Additionally, a lack of understanding regarding all potentially pathogenic variants carried by ALS patients may limit their enrolment in precision medicine-based clinical trials.

In contrast to the risk associations, our results suggested that the association between oligogenicity and clinical outcomes of ALS remains unclear and might not involve the same genes. The only association between ALS age of onset and carrying more than one rare variant in a known ALS gene was observed when considering missense variants and PTVs with an MAF < 0.01, and the *C9orf72* repeat expansion (p = 0.042). Yet, this association was only nominally significant, which — in addition to the absence of further oligogenic associations with ALS age of onset — results in a lack of clarity regarding the validity of this relationship. We only observed an association between singleton carriers and survival period when the *C9orf72* repeat expansion was included in the analysis, which may be explained by the *C9orf72* pathogenic repeat expansion being associated with a decreased survival period individually^43,44^. Overall, consistent with recent reports^11,45,46^, these results suggest the genetic architecture underlying ALS risk is decoupled from that underlying survival. Moreover, considering that every modification of risk is expected to correspond to an effect on age of onset^16,47^, our results suggest that this relationship might only explain a limited proportion of the age of onset variability in ALS and the presence of concurring independent mechanisms with large effect is possible.

Oligogenic events involving the *C9orf72* repeat expansion were particularly frequent among the individuals with ALS (1.48%), the statistical power from which may contribute to our observations of the repeat’s contribution to clinical outcomes described above. As one of the most commonly inherited forms of ALS in Europeans^48^, we sought to examine whether the oligogenic effect was primarily driven by this repeat expansion. Excluding *C9orf72,* our risk assessments confirmed that oligogenic events involving missense variants and PTVs, both in the presence and absence of the *ATXN2* repeat expansion, conferred significant risk to ALS. While a previous assessment of oligogenicity involving *C9orf72* suggested the repeat expansion was sufficient to cause ALS alone^49^, our results suggested an increased risk from oligogenic events involving *C9orf72* in comparison to *C9orf72* singleton events. Oligogenic events could help explain the recent incomplete penetrance estimates of *C9orf72* expansions^50^. Further, Ciura et al identified pathways by which the *C9orf72* and *ATXN2* pathogenic repeat expansions may genetically interact, suggesting an actual biological impact of oligogenicity involving *C9orf72* in ALS pathogenesis^51^. Additional functional analyses will be required to determine how variants in multiple known ALS genes may synergistically induce pathology.

Many other known ALS genes were also more commonly observed oligogenic events carried by individuals with ALS than controls. Oligogenic events in the genes *CHMP2B*, *PFN1*, *SOD1*, and *UBQLN2* were entirely unique to individuals with ALS in the discovery subset (absent in controls). Oligogenic events in the genes *DNAJC7*, *FUS*, *HNRNPA1*, *TUB4A4*, and *VCP* or the *C9orf72* repeat expansion were only each observed in one control (0.055%). Of these ten genes, seven are established ALS genes — referring to those that have been classified as having a ‘definitive’ relationship with ALS according to the ClinGen ALS GCEP. While it could be proposed that oligogenic events involving established ALS genes are driving the observed risk associations, a large proportion of individuals with ALS carried oligogenic events involving only ALS-associated genes — referring to genes that have not been defined as having a definitive relationship with ALS according to the ClinGen ALS GCEP — or involving at least one variant in an established ALS gene and at least one variant in an ALS-associated gene (2.8%). We suspect that these oligogenic events involving variants in ALS-associated genes may encompass a large subset of cases in which only a singleton variant may not have contributed enough risk to drive disease onset. Yet how the two variants within each identified oligogenic event interact remains unknown, and it is possible that some events may represent cases of genetic modification — encompassing a variant driving disease risk in combination with a variant modifying disease presentation — as has been observed for oligogenic cases of complex neuropathy and retinal degeneration, among other diseases^8,52–56^.

Collectively, our results reveal that oligogenic events contribute significant risk to ALS, both in the presence and absence of variants in the most well-established ALS genes, such as *C9orf72*. Moreover, the observed lack of influence of oligogenicity on survival of ALS supports the recent hypothesis of decoupling between mechanisms underlying the risk of ALS and its progression. Although our study represents the largest systematic analysis of oligogenicity to date, even greater sample sizes and variant effect studies are required to determine the exact consequences of carrying multiple ALS-associated variants on disease progression and outcomes. Nevertheless, our findings confirm that oligogenic events are relevant in ALS, which may be of particular importance when the variants involved have uncertain pathogenic significance or are observed in genes with probable ALS associations. The potential implications of these variants on ALS clinical correlates and molecular pathology warrant further exploration. In the age of stratified medication and gene therapy, implicating oligogenicity in a relevant proportion of ALS patients supports the need for a complete genetic profile for accurate genetic counselling and the correct choice of therapy in all ALS patients.

## Data availability

Individual whole-genome sequencing data are available and can be requested through Project MinE (https://www.projectmine.com/research/data-sharing/). A data access committee controls access to raw data, ensuring a FAIR data setup (https://www.datafairport.org). Details on the frequencies and gene burden test results are available on the ProjectMinE databrowser ^29^ (http://databrowser.projectmine.com).

## Acknowledgments

Samples used in this research were in part obtained from the UK National DNA Bank for MND Research, funded by the MND Association and the Wellcome Trust. Part of the samples were obtained from The Project MinE and MND centres internationally. We thank people with MND and their families for their participation in this project. The authors acknowledge use of the King’s Computational Research, Engineering and Technology Environment (CREATE) (https://create.kcl.ac.uk), which is delivered in partnership with the National Institute for Health and Care Research (NIHR) Biomedical Research Centres at South London and Maudsley and Guy’s and St. Thomas’ NHS Foundation Trusts, and part-funded by capital equipment grants from the Maudsley Charity (award 980) and Guy’s and St. Thomas’ Charity (TR130505). We also acknowledge Health Data Research UK, which is funded by the UK Medical Research Council, Engineering and Physical Sciences Research Council, Economic and Social Research Council, Department of Health and Social Care (United Kingdom), Chief Scientist Office of the Scottish Government Health and Social Care Directorates, Health and Social Care Research and Development Division (Welsh Government), Public Health Agency (Northern Ireland), British Heart Foundation and Wellcome Trust.

---

**Supplemental Table 1.**
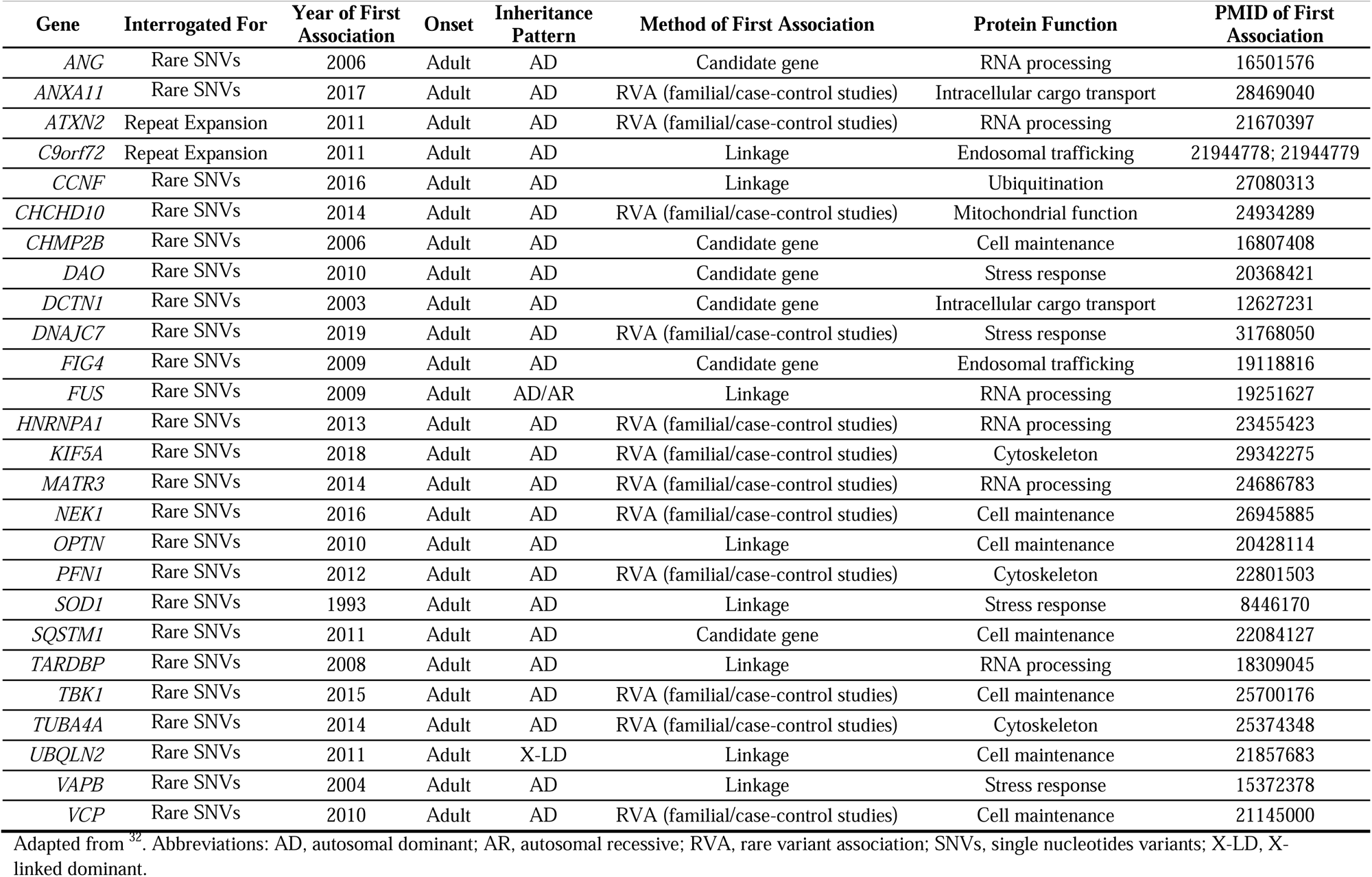
Twenty-six known amyotrophic lateral sclerosis (ALS) genes interrogated for rare variants or repeat expansions in the genome sequencing datasets of the Project MinE ALS Sequencing Consortium.

| Gene | Interrogated For | Year of First Association | Onset | Inheritance Pattern | Method of First Association | Protein Function | PMID of First Association |
| --- | --- | --- | --- | --- | --- | --- | --- |
| *ANG* | Rare SNVs | 2006 | Adult | AD | Candidate gene | RNA processing | 16501576 |
| *ANXA11* | Rare SNVs | 2017 | Adult | AD | RVA (familial/case-control studies) | Intracellular cargo transport | 28469040 |
| *ATXN2* | Repeat Expansion | 2011 | Adult | AD | RVA (familial/case-control studies) | RNA processing | 21670397 |
| *C9orf72* | Repeat Expansion | 2011 | Adult | AD | Linkage | Endosomal trafficking | 21944778; 21944779 |
| *CCNF* | Rare SNVs | 2016 | Adult | AD | Linkage | Ubiquitination | 27080313 |
| *CHCHD10* | Rare SNVs | 2014 | Adult | AD | RVA (familial/case-control studies) | Mitochondrial function | 24934289 |
| *CHMP2B* | Rare SNVs | 2006 | Adult | AD | Candidate gene | Cell maintenance | 16807408 |
| *DAO* | Rare SNVs | 2010 | Adult | AD | Candidate gene | Stress response | 20368421 |
| *DCTN1* | Rare SNVs | 2003 | Adult | AD | Candidate gene | Intracellular cargo transport | 12627231 |
| *DNAJC7* | Rare SNVs | 2019 | Adult | AD | RVA (familial/case-control studies) | Stress response | 31768050 |
| *FIG4* | Rare SNVs | 2009 | Adult | AD | Candidate gene | Endosomal trafficking | 19118816 |
| *FUS* | Rare SNVs | 2009 | Adult | AD/AR | Linkage | RNA processing | 19251627 |
| *HNRNPA1* | Rare SNVs | 2013 | Adult | AD | RVA (familial/case-control studies) | RNA processing | 23455423 |
| *KIF5A* | Rare SNVs | 2018 | Adult | AD | RVA (familial/case-control studies) | Cytoskeleton | 29342275 |
| *MATR3* | Rare SNVs | 2014 | Adult | AD | RVA (familial/case-control studies) | RNA processing | 24686783 |
| *NEK1* | Rare SNVs | 2016 | Adult | AD | RVA (familial/case-control studies) | Cell maintenance | 26945885 |
| *OPTN* | Rare SNVs | 2010 | Adult | AD | Linkage | Cell maintenance | 20428114 |
| *PFN1* | Rare SNVs | 2012 | Adult | AD | RVA (familial/case-control studies) | Cytoskeleton | 22801503 |
| *SOD1* | Rare SNVs | 1993 | Adult | AD | Linkage | Stress response | 8446170 |
| *SQSTM1* | Rare SNVs | 2011 | Adult | AD | Candidate gene | Cell maintenance | 22084127 |
| *TARDBP* | Rare SNVs | 2008 | Adult | AD | Linkage | RNA processing | 18309045 |
| *TBK1* | Rare SNVs | 2015 | Adult | AD | RVA (familial/case-control studies) | Cell maintenance | 25700176 |
| *TUBA4A* | Rare SNVs | 2014 | Adult | AD | RVA (familial/case-control studies) | Cytoskeleton | 25374348 |
| *UBQLN2* | Rare SNVs | 2011 | Adult | X-LD | Linkage | Cell maintenance | 21857683 |
| *VAPB* | Rare SNVs | 2004 | Adult | AD | Linkage | Stress response | 15372378 |
| *VCP* | Rare SNVs | 2010 | Adult | AD | RVA (familial/case-control studies) | Cell maintenance | 21145000 |

Adapted from<sup>33</sup>. Abbreviations: AD, autosomal dominant; AR, autosomal recessive; RVA, rare variant association; SNVs, single nucleotides variants; X-LD, X-linked dominant.

**Supplemental Figure 1.**
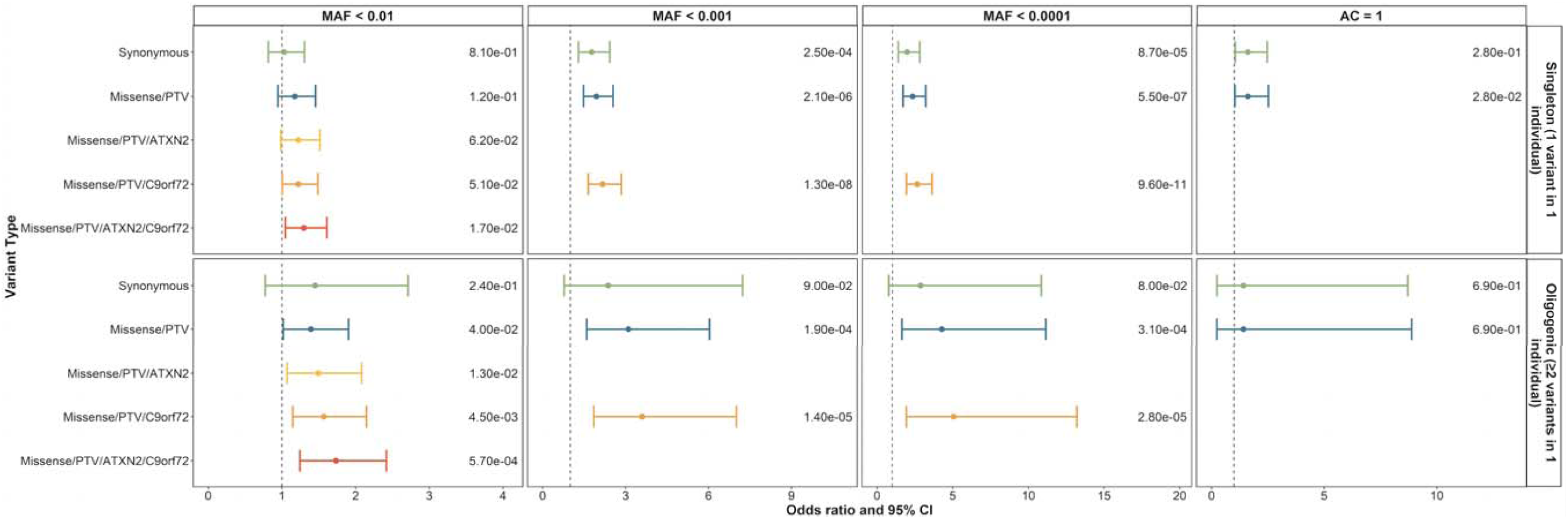
Replication of singleton and oligogenic enrichment of rare variants with various MAF in known ALS genes in individuals with ALS compared to controls. Enrichment of carriers of rare variants in one known ALS gene and carriers of rare variants in two or more known ALS genes was compared to non-carriers in individuals with ALS compared to controls. Enrichment analyses were performed using logistic regression in a replication (individuals with ALS = 2057, controls = 513) subset of the Project MinE ALS sequencing consortium dataset, including sex, 10 ancestry defining principal components, and total variant count (total genetic load) as covariates. *ATXN2* and *C9orf72* refer to a pathogenic repeat expansion being identified in the respective gene using ExpansionHunter. Minor allele frequencies were obtained from the GnomAD v2.1.1 non-neurological dataset. AC = 1 refers to variants absent from the GnomAD v2.1.1 non-neurological dataset and only a single observation in the analyzed dataset. Logistic regression generated p-values are displayed on the right side of each plot. Abbreviations: AC, allele count; CI, confidence interval; MAF, minor allele frequency; PTV, protein truncating variant.

**Supplemental Figure 2.**
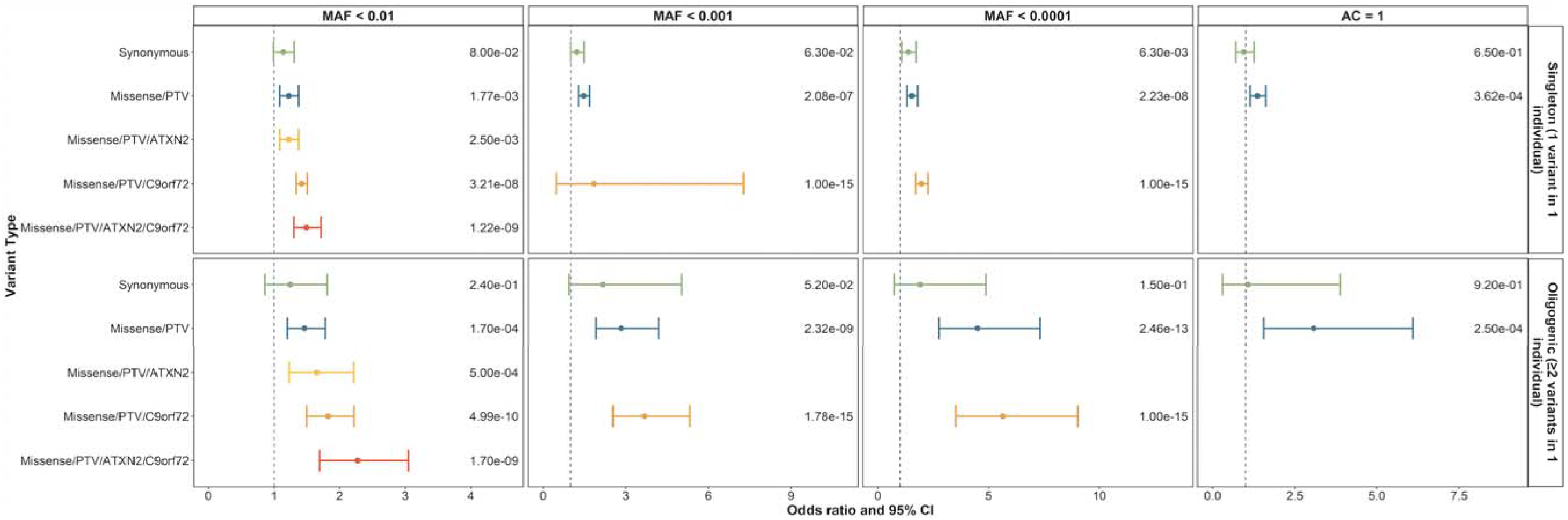
Singleton and oligogenic enrichment of rare variants with various MAF in known ALS genes in individuals with ALS compared to controls. Enrichment of carriers of rare variants in one known ALS gene and carriers of rare variants in two or more known ALS genes was compared to non-carriers in individuals with ALS compared to controls. Enrichment analyses were performed using logistic regression in a discovery (individuals with ALS = 4299, controls = 1815) subset of the Project MinE ALS sequencing consortium dataset, including sex, 10 ancestry defining principal components, and total variant count (total genetic load) as covariates. *ATXN2* and *C9orf72* refer to a pathogenic repeat expansion being identified in the respective gene using ExpansionHunter. Minor allele frequencies were obtained from the GnomAD v2.1.1 non-neurological dataset. AC = 1 refers to variants absent from the GnomAD v2.1.1 non-neurological dataset and only a single observation in the analyzed dataset. Logistic regression generated p-values are displayed on the right side of each plot. Abbreviations: AC, allele count; CI, confidence interval; MAF, minor allele frequency; PTV, protein truncating variant.

**Supplemental Figure 3.**
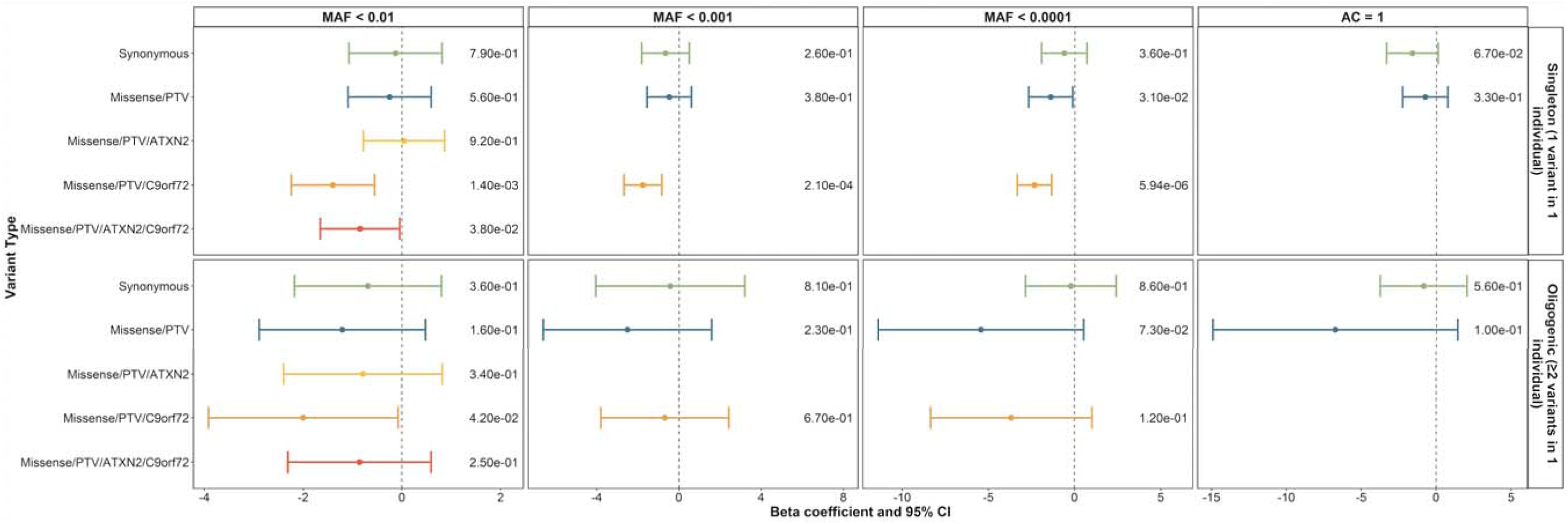
Influence of singleton and oligogenic enrichment of rare variants with various MAF in known ALS genes to ALS age of onset. The influence of carrying a rare variant of various minor allele frequencies in a single known known ALS gene was compared to the influence of carrying rare variants in two or more known ALS genes on ALS age of onset in the discovery cohort of the Project MinE ALS sequencing consortium (individuals with ALS = 4299). Enrichment analyses were performed using linear regression including sex, site of onset, 10 ancestry defining principal components, and total variant count (total genetic load) as covariates. *ATXN2* and *C9orf72* refer to a pathogenic repeat expansion being identified in the respective gene using ExpansionHunter. Minor allele frequencies were obtained from the GnomAD v2.1.1 non-neurological dataset. AC = 1 refers to variants absent from the GnomAD v2.1.1 non-neurological dataset and only a single observation in the analyzed dataset. Linear regression generated p-values are displayed on the right side of each plot. Abbreviations: AC, allele count; CI, confidence interval; MAF, minor allele frequency; PTV, protein truncating variant.

**Supplemental Figure 4.**
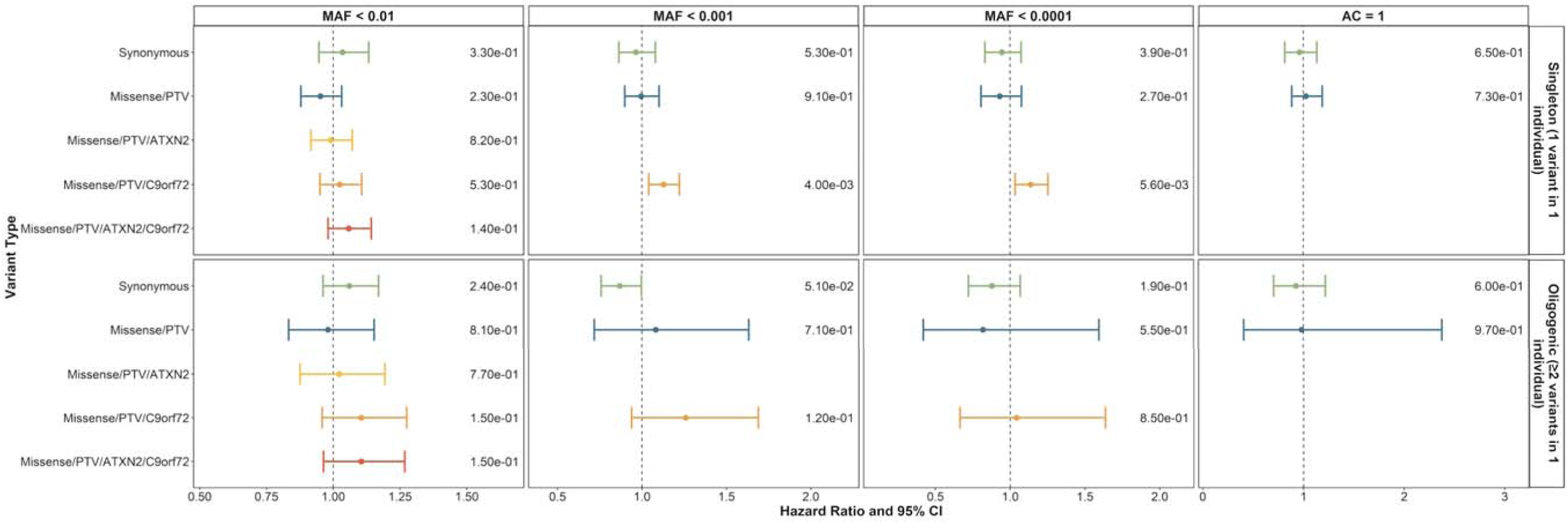
Influence of singleton and oligogenic enrichment of rare variants with various MAF in known ALS genes to ALS survival period. The influence of carrying a rare variant of various minor allele frequencies in a single known ALS gene was compared to the influence of carrying rare variants in two or more known ALS genes on ALS survival period in the discovery cohort of the Project MinE ALS sequencing consortium (individuals with ALS = 4299). Enrichment analyses were performed using a Cox proportional-hazards model including sex, site of onset, 10 ancestry defining principal components, and total variant count (total genetic load) as covariates. Synonymous, missense, and protein truncating variants (PTVs) were identified in 24 known ALS genes using whole genome sequencing. *ATXN2* and *C9orf72* refer to a pathogenic repeat expansion being identified in the respective gene using ExpansionHunter. Minor allele frequencies were obtained from the GnomAD v2.1.1 non-neurological dataset. AC = 1 refers to variants absent from the GnomAD v2.1.1 non-neurological dataset and only a single observation in the analyzed dataset. Survival period was defined as years from diagnosis to death, or years from diagnosis to last follow-up, as appropriate. Cox proportional-hazards model generated p-values are displayed on the right side of each plot. Abbreviations: AC, allele count; CI, confidence interval; MAF, minor allele frequency; PTV, protein truncating variant.

**Supplemental Figure 5.**
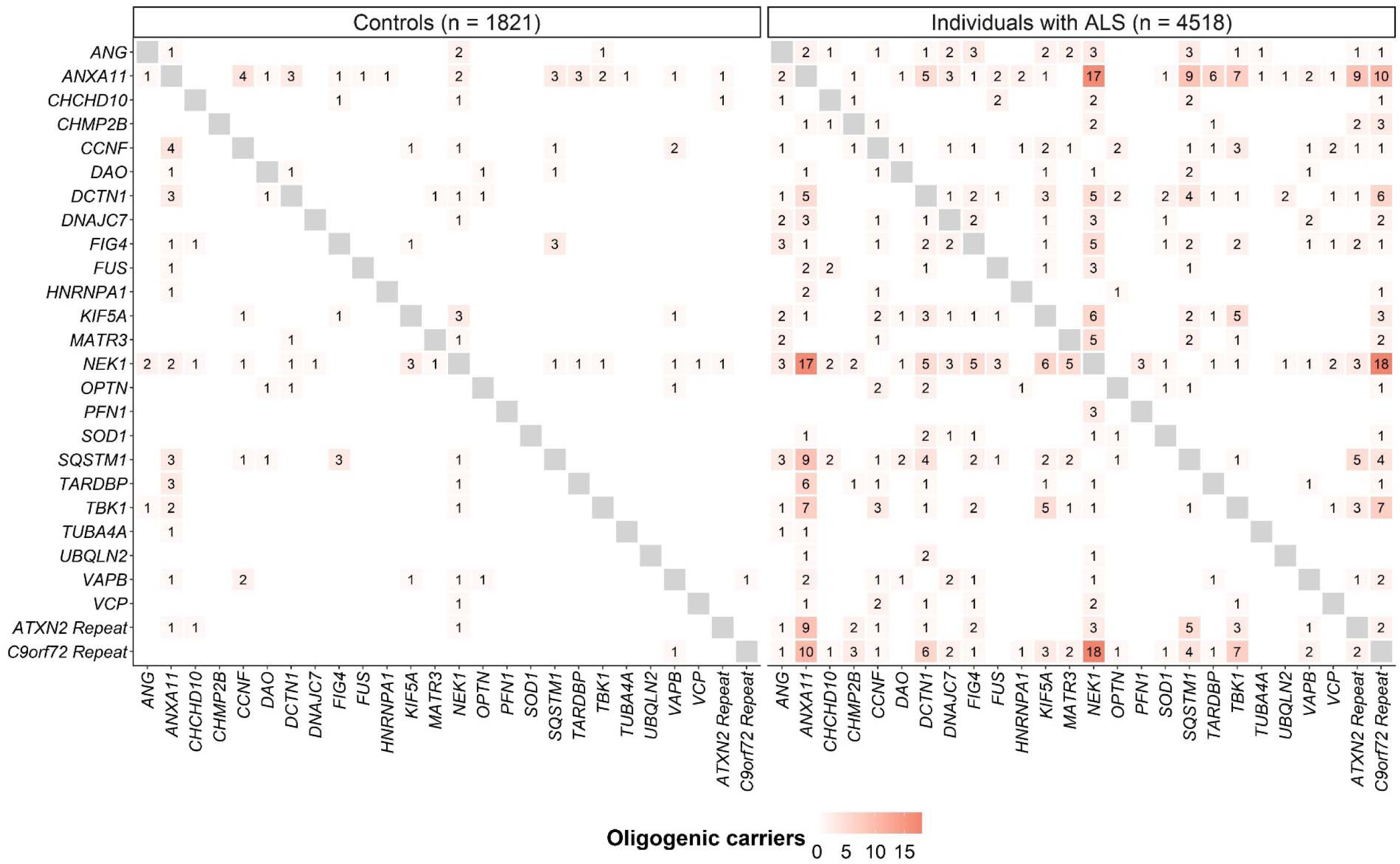
Comparison of the number of individuals with ALS (n = 4,518) and controls (n = 1,821) that were oligogenic rare variant (MAF < 0.01) carriers. The gene matrices display the number of individuals with ALS and controls from the Project MinE discovery cohort carrying at least one rare variant in each gene encompassed in the respective column and row.

**Supplemental Figure 6.**
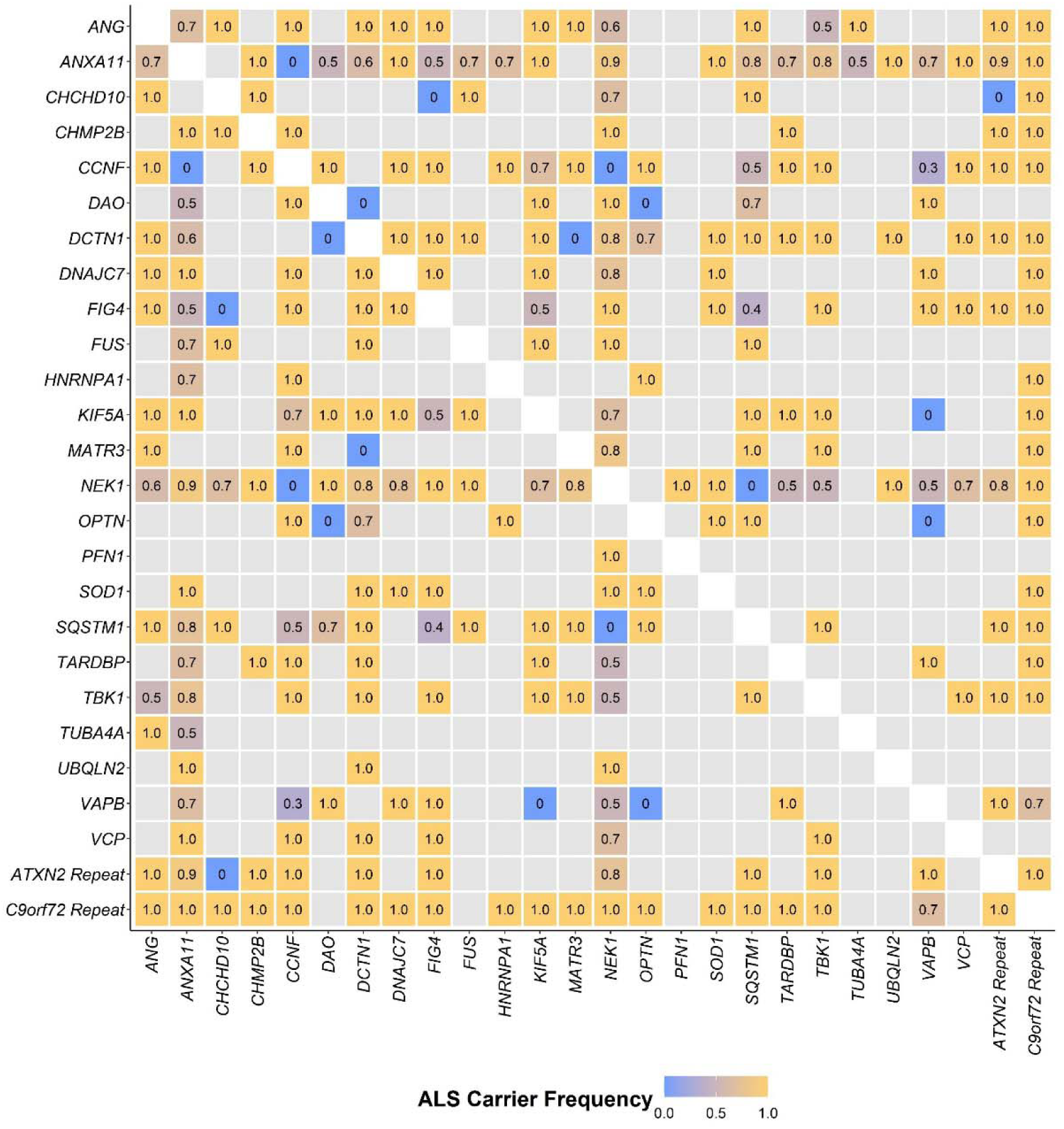
Frequency of oligogenic rare variant (MAF < 0.01) carriers that had ALS. The gene matrix displays the ALS carrier frequency with at least one rare variant in each gene encompassed in the respective column and row based on the discovery cohort of the Project MinE ALS sequencing consortium (individuals with ALS = 4299, controls = 1815). ALS carrier frequency was calculated by dividing the number of individuals with ALS carrying rare variants in the specific gene combination with the total number of samples from the Project MinE discovery cohort carrying rare variants in the specific gene combination.

## References

1. Ben-Mahmoud A, Gupta V, Kim C-H, Layman LC, Kim H-G. Digenic or oligogenic mutations in presumed monogenic disorders: A review. J Genet Med. 2023;20(1):15–24. doi:10.5734/JGM.2023.20.1.15

2. Katsanis N, Ansley SJ, Badano JL, et al. Triallelic inheritance in Bardet-Biedl syndrome, a Mendelian recessive disorder. Science. Sep 21 2001;293(5538):2256–9. doi:10.1126/science.1063525

3. Leitch CC, Zaghloul NA, Davis EE, et al. Hypomorphic mutations in syndromic encephalocele genes are associated with Bardet-Biedl syndrome. Nat Genet. Apr 2008;40(4):443–8. doi:10.1038/ng.97

4. Perea-Romero I, Solarat C, Blanco-Kelly F, et al. Allelic overload and its clinical modifier effect in Bardet-Biedl syndrome. NPJ Genom Med. Jul 14 2022;7(1):41. doi:10.1038/s41525-022-00311-2

5. Giudicessi JR, Wilde AAM, Ackerman MJ. The genetic architecture of long QT syndrome: A critical reappraisal. Trends Cardiovasc Med. Oct 2018;28(7):453–464. doi:10.1016/j.tcm.2018.03.003

6. Gifford CA, Ranade SS, Samarakoon R, et al. Oligogenic inheritance of a human heart disease involving a genetic modifier. Science. May 31 2019;364(6443):865–870. doi:10.1126/science.aat5056

7. Kousi M, Soylemez O, Ozanturk A, et al. Evidence for secondary-variant genetic burden and non-random distribution across biological modules in a recessive ciliopathy. Nat Genet. Nov 2020;52(11):1145–1150. doi:10.1038/s41588-020-0707-1

8. Gonzaga-Jauregui C, Harel T, Gambin T, et al. Exome Sequence Analysis Suggests that Genetic Burden Contributes to Phenotypic Variability and Complex Neuropathy. Cell Rep. Aug 18 2015;12(7):1169–83. doi:10.1016/j.celrep.2015.07.023

9. Farhan SMK, Howrigan DP, Abbott LE, et al. Exome sequencing in amyotrophic lateral sclerosis implicates a novel gene, DNAJC7, encoding a heat-shock protein. Nat Neurosci. Dec 2019;22(12):1966–1974. doi:10.1038/s41593-019-0530-0

10. Zhang S, Cooper-Knock J, Weimer AK, et al. Genome-wide identification of the genetic basis of amyotrophic lateral sclerosis. Neuron. Mar 16 2022;110(6):992–1008 e11. doi:10.1016/j.neuron.2021.12.019

11. van Rheenen W, van der Spek RAA, Bakker MK, et al. Common and rare variant association analyses in amyotrophic lateral sclerosis identify 15 risk loci with distinct genetic architectures and neuron-specific biology. Nat Genet. Dec 2021;53(12):1636–1648. doi:10.1038/s41588-021-00973-1

12. van Rheenen W, Shatunov A, Dekker AM, et al. Genome-wide association analyses identify new risk variants and the genetic architecture of amyotrophic lateral sclerosis. Nat Genet. Sep 2016;48(9):1043–8. doi:10.1038/ng.3622

13. Cooper-Knock J, Zhang S, Kenna KP, et al. Rare Variant Burden Analysis within Enhancers Identifies CAV1 as an ALS Risk Gene. Cell Rep. Dec 1 2020;33(9):108456. doi:10.1016/j.celrep.2020.108456

14. Smith BN, Topp SD, Fallini C, et al. Mutations in the vesicular trafficking protein annexin A11 are associated with amyotrophic lateral sclerosis. Sci Transl Med. May 3 2017;9(388)doi:10.1126/scitranslmed.aad9157

15. Iacoangeli A, Lin T, Al Khleifat A, et al. Genome-wide Meta-analysis Finds the ACSL5-ZDHHC6 Locus Is Associated with ALS and Links Weight Loss to the Disease Genetics. Cell Rep. Oct 27 2020;33(4):108323. doi:10.1016/j.celrep.2020.108323

16. Mehta PR, Iacoangeli A, Opie-Martin S, et al. The impact of age on genetic testing decisions in amyotrophic lateral sclerosis. Brain. Dec 19 2022;145(12):4440–4447. doi:10.1093/brain/awac279

17. van Blitterswijk M, van Es MA, Hennekam EA, et al. Evidence for an oligogenic basis of amyotrophic lateral sclerosis. Hum Mol Genet. Sep 1 2012;21(17):3776–84. doi:10.1093/hmg/dds199

18. McCann EP, Henden L, Fifita JA, et al. Evidence for polygenic and oligogenic basis of Australian sporadic amyotrophic lateral sclerosis. J Med Genet. May 14 2020;doi:10.1136/jmedgenet-2020-106866

19. Al-Chalabi A, Calvo A, Chio A, et al. Analysis of amyotrophic lateral sclerosis as a multistep process: a population-based modelling study. Lancet Neurol. Nov 2014;13(11):1108–1113. doi:10.1016/S1474-4422(14)70219-4

20. Chio A, Mazzini L, D’Alfonso S, et al. The multistep hypothesis of ALS revisited: The role of genetic mutations. Neurology. Aug 14 2018;91(7):e635–e642. doi:10.1212/WNL.0000000000005996

21. Cady J, Allred P, Bali T, et al. Amyotrophic lateral sclerosis onset is influenced by the burden of rare variants in known amyotrophic lateral sclerosis genes. Ann Neurol. Jan 2015;77(1):100–13. doi:10.1002/ana.24306

22. Kuuluvainen L, Kaivola K, Monkare S, et al. Oligogenic basis of sporadic ALS: The example of SOD1 p.Ala90Val mutation. Neurol Genet. Jun 2019;5(3):e335. doi:10.1212/NXG.0000000000000335

23. Zhang H, Cai W, Chen S, et al. Screening for possible oligogenic pathogenesis in Chinese sporadic ALS patients. Amyotroph Lateral Scler Frontotemporal Degener. Aug 2018;19(5-6):419–425. doi:10.1080/21678421.2018.1432659

24. Farhan SMK, Gendron TF, Petrucelli L, Hegele RA, Strong MJ. OPTN p.Met468Arg and ATXN2 intermediate length polyQ extension in families with C9orf72 mediated amyotrophic lateral sclerosis and frontotemporal dementia. Am J Med Genet B Neuropsychiatr Genet. Jan 2018;177(1):75–85. doi:10.1002/ajmg.b.32606

25. Farhan SMK, Gendron TF, Petrucelli L, Hegele RA, Strong MJ. ARHGEF28 p.Lys280Metfs40Ter in an amyotrophic lateral sclerosis family with a C9orf72 expansion. Neurol Genet. Oct 2017;3(5):e190. doi:10.1212/NXG.0000000000000190

26. Volkening K, Farhan SMK, Kao J, et al. Evidence of synergism among three genetic variants in a patient with LMNA-related lipodystrophy and amyotrophic lateral sclerosis leading to a remarkable nuclear phenotype. Mol Cell Biochem. Jul 2021;476(7):2633–2650. doi:10.1007/s11010-021-04103-7

27. Van Daele SH, Moisse M, van Vugt J, et al. Genetic variability in sporadic amyotrophic lateral sclerosis. Brain. Sep 1 2023;146(9):3760-3769. doi:10.1093/brain/awad120

28. Project Min EALSSC. Project MinE: study design and pilot analyses of a large-scale whole-genome sequencing study in amyotrophic lateral sclerosis. Eur J Hum Genet. Oct 2018;26(10):1537–1546. doi:10.1038/s41431-018-0177-4

29. van der Spek RAA, van Rheenen W, Pulit SL, et al. The project MinE databrowser: bringing large-scale whole-genome sequencing in ALS to researchers and the public. Amyotroph Lateral Scler Frontotemporal Degener. Aug 2019;20(5-6):432–440. doi:10.1080/21678421.2019.1606244

30. McLaren W, Gil L, Hunt SE, et al. The Ensembl Variant Effect Predictor. Genome Biol. Jun 6 2016;17(1):122. doi:10.1186/s13059-016-0974-4

31. Wang K, Li M, Hakonarson H. ANNOVAR: functional annotation of genetic variants from high-throughput sequencing data. Nucleic Acids Res. Sep 2010;38(16):e164. doi:10.1093/nar/gkq603

32. Karczewski KJ, Francioli LC, Tiao G, et al. The mutational constraint spectrum quantified from variation in 141,456 humans. Nature. May 2020;581(7809):434–443. doi:10.1038/s41586-020-2308-7

33. Dilliott AA, Rouleau GA, Iqbal S, Farhan SMK. Characterizing proteomic and transcriptomic features of missense variants in amyotrophic lateral sclerosis genes. medRxiv. 2022:2022.12.21.22283728. doi:10.1101/2022.12.21.22283728

34. Dolzhenko E, Deshpande V, Schlesinger F, et al. ExpansionHunter: a sequence-graph-based tool to analyze variation in short tandem repeat regions. Bioinformatics. Nov 1 2019;35(22):4754–4756. doi:10.1093/bioinformatics/btz431

35. Dolzhenko E, van Vugt J, Shaw RJ, et al. Detection of long repeat expansions from PCR-free whole-genome sequence data. Genome Res. Nov 2017;27(11):1895–1903. doi:10.1101/gr.225672.117

36. Iacoangeli A, Al Khleifat A, Jones AR, et al. C9orf72 intermediate expansions of 24-30 repeats are associated with ALS. Acta Neuropathol Commun. Jul 17 2019;7(1):115. doi:10.1186/s40478-019-0724-4

37. Tazelaar GHP, Boeynaems S, De Decker M, et al. ATXN1 repeat expansions confer risk for amyotrophic lateral sclerosis and contribute to TDP-43 mislocalization. Brain Commun. 2020;2(2):fcaa064. doi:10.1093/braincomms/fcaa064

38. DeJesus-Hernandez M, Mackenzie IR, Boeve BF, et al. Expanded GGGGCC hexanucleotide repeat in noncoding region of C9ORF72 causes chromosome 9p-linked FTD and ALS. Neuron. Oct 20 2011;72(2):245–56. doi:10.1016/j.neuron.2011.09.011

39. Renton AE, Majounie E, Waite A, et al. A hexanucleotide repeat expansion in C9ORF72 is the cause of chromosome 9p21-linked ALS-FTD. Neuron. Oct 20 2011;72(2):257–68. doi:10.1016/j.neuron.2011.09.010

40. Daoud H, Belzil V, Martins S, et al. Association of long ATXN2 CAG repeat sizes with increased risk of amyotrophic lateral sclerosis. Arch Neurol. Jun 2011;68(6):739–42. doi:10.1001/archneurol.2011.111

41. R: A Language and Environment for Statistical Computing. R Foundation for Statistical Computing; 2021. https://www.R-project.org

42. Wickham H. Ggplot2: Elegant Graphics for Data Analysis. 2nd Edition ed. Springer; 2009.

43. Umoh ME, Fournier C, Li Y, et al. Comparative analysis of C9orf72 and sporadic disease in an ALS clinic population. Neurology. Sep 6 2016;87(10):1024–30. doi:10.1212/WNL.0000000000003067

44. Su WM, Gu XJ, Duan QQ, et al. Genetic factors for survival in amyotrophic lateral sclerosis: an integrated approach combining a systematic review, pairwise and network meta-analysis. BMC Med. Jun 27 2022;20(1):209. doi:10.1186/s12916-022-02411-3

45. Opie-Martin S, Iacoangeli A, Topp SD, et al. The SOD1-mediated ALS phenotype shows a decoupling between age of symptom onset and disease duration. Nat Commun. Nov 12 2022;13(1):6901. doi:10.1038/s41467-022-34620-y

46. Kalia M, Miotto M, Ness D, et al. Molecular dynamics analysis of Superoxide Dismutase 1 mutations suggests decoupling between mechanisms underlying ALS onset and progression. Computational and Structural Biotechnology Journal. 2023;doi:10.1016/j.csbj.2023.09.016

47. Mehta PR, Jones AR, Opie-Martin S, et al. Younger age of onset in familial amyotrophic lateral sclerosis is a result of pathogenic gene variants, rather than ascertainment bias. J Neurol Neurosurg Psychiatry. Mar 2019;90(3):268–271. doi:10.1136/jnnp-2018-319089

48. Smith BN, Newhouse S, Shatunov A, et al. The C9ORF72 expansion mutation is a common cause of ALS+/-FTD in Europe and has a single founder. Eur J Hum Genet. Jan 2013;21(1):102–8. doi:10.1038/ejhg.2012.98

49. Ross JP, Leblond CS, Laurent SB, et al. Oligogenicity, C9orf72 expansion, and variant severity in ALS. Neurogenetics. Jul 2020;21(3):227–242. doi:10.1007/s10048-020-00612-7

50. Spargo TP, Opie-Martin S, Bowles H, Lewis CM, Iacoangeli A, Al-Chalabi A. Calculating variant penetrance from family history of disease and average family size in population-scale data. Genome Med. Dec 15 2022;14(1):141. doi:10.1186/s13073-022-01142-7

51. Ciura S, Sellier C, Campanari ML, Charlet-Berguerand N, Kabashi E. The most prevalent genetic cause of ALS-FTD, C9orf72 synergizes the toxicity of ATXN2 intermediate polyglutamine repeats through the autophagy pathway. Autophagy. Aug 2 2016;12(8):1406–8. doi:10.1080/15548627.2016.1189070

52. Louie CM, Caridi G, Lopes VS, et al. AHI1 is required for photoreceptor outer segment development and is a modifier for retinal degeneration in nephronophthisis. Nat Genet. Feb 2010;42(2):175–80. doi:10.1038/ng.519

53. Smemo S, Tena JJ, Kim KH, et al. Obesity-associated variants within FTO form long-range functional connections with IRX3. Nature. Mar 20 2014;507(7492):371–5. doi:10.1038/nature13138

54. Darrah R, McKone E, O’Connor C, et al. EDNRA variants associate with smooth muscle mRNA levels, cell proliferation rates, and cystic fibrosis pulmonary disease severity. Physiol Genomics. Mar 3 2010;41(1):71–7. doi:10.1152/physiolgenomics.00185.2009

55. Hillian AD, Londono D, Dunn JM, et al. Modulation of cystic fibrosis lung disease by variants in interleukin-8. Genes Immun. Sep 2008;9(6):501–8. doi:10.1038/gene.2008.42

56. Kousi M, Katsanis N. Genetic modifiers and oligogenic inheritance. Cold Spring Harb Perspect Med. Jun 1 2015;5(6)doi:10.1101/cshperspect.a017145

